# ‘Grinning and bearing it’ - A mixed methods approach to explore animal-related injuries in UK and Irish Veterinary Students

**DOI:** 10.64898/2025.12.19.25342672

**Authors:** Tamzin Furtado, Lois Kennedy, Gina Pinchbeck, John S P Tulloch

## Abstract

**Background:** While veterinary surgeons are known to have particularly high rates of injury compared to other sectors, little is known about rates of injury among veterinary students. This study aims to understand animal-related injury rates, injury context and mechanisms, attitudes to reporting injuries, and behaviour change among UK and Irish veterinary students.

**Methods:** A survey was distributed to students across all veterinary schools operating in the UK and Ireland in 2021. Questions explored participants experience of injury through asking about their most recent and most severe injuries via quantitative and free-text questions. Data were analysed using descriptive statistics, logistic regression, and qualitative content analysis.

**Results:** 533 responses were included in the analyses. Overall, 47.5% of students reported having been injured by an animal during the veterinary degree, 35.5% of students reported being injured within the last 12 months. Most recent injuries were caused by companion animals (38.0%), livestock (37.6%), and equids (23.5%). For their most severe injuries, 48.7% involved livestock, 28.7% companion animals, and 22.1% equids. The content analysis highlighted that students normalised injuries and infrequently reported injuries to the university. It was very rare for students to take time off from their studies or placements, due to course pressures.

**Conclusions:** These findings reflect concerningly high levels of injury, which are being under-reported and reflect a culture of injury acceptance and expectation among students. Veterinary schools should consider lessons learnt in other work environments which have been successful in changing safety culture.

## Introduction

The veterinary profession has some of the highest levels of non-fatal injuries. In 2023 the US Bureau of Labor Statistics recorded that ‘Veterinary services’ had the second highest non-fatal injury incidence rate out of any industry, 10.6 injuries per 100 full-time equivalents, this amounts to around 38,000 cases annually (1), more than four times the national incidence rate of 2.5.

Data regarding the incidence of non-fatal injuries within the British and Irish veterinary workforces is challenging, as the industry is so small that data are often not presented. Survey data has suggested that being an equine veterinarian was one of the most hazardous civilian professions (2), and that 58.3% of companion animal veterinarians (3), 48.6% of equine veterinarians, and 62.5% of large animal veterinarians are injured at work annually (4). These injuries are predominately animal-related (3-10). US data suggests that animal-related injuries make up 49.3% of all injuries that result in days away from work, a further 23.2% were due to falls, slips or trips (1).

Between 25-33% of British equine veterinarians attend a hospital when they get injured (2, 4). Accident books from United Kingdom (UK) veterinary schools showed that 16% of staff injured by a horse required hospital attendance (5). Horse-related injuries were predominately caused by kicks whilst performing a clinical exam, in particular distal limb examinations (4). Whilst cattle-related injuries were mainly kicks and crushes whilst performing a clinical exam, with 21-40% requiring hospital attendance (4, 5). Dogs and cats generally cause a higher frequency of injuries, mainly through bites and scratches (3, 5, 6). Other work-related injuries in the veterinary sector include needlestick and sharp injuries, traffic accidents, and ergonomic injuries (3, 6, 11, 12). The top three most prevalent injury types within the sector which result in days away from work, restricted activity, or job transfer are; puncture wound (48.6%), fractures (12.7%) and sprains (9.2%) (1).

There is emerging evidence that the profession has a complex relationship with work-related injuries, with a culture that minimises the risk and consequences of injury and thus promotes injury acceptance as “just part of the job” (3, 13, 14), in particular the risk posed by animals (15). Many within the profession only identify something as a work-related injury if it limits their ability to perform their job or that they have to attend a hospital (14). Given this context it is important to understand whether these same injuries are prevalent for veterinary students, and whether the context and culture around them is similar. If they are similar then veterinary schools provide the perfect place to challenge the cultural norms, and to develop intervention and education programmes to minimise injury risk.

Several studies have provided insight into veterinary student physical injury; for example an exploration of injury reports in UK veterinary schools over a ten year period found that students had lower levels of injuries than staff, and that injuries were predominantly associated with cats and dogs (5). However, since students are known to under-report zoonotic infections it is likely that they also under-report injuries, and that these figures may not be representative (16). In Australia, the most prevalent cause of injury to veterinary students were sharps injuries (40.0%), animal bites (39.6%), and being rammed/pushed over by a large animal (6). The rates of injuries were higher than that of veterinarians or veterinary nurses. In the UK and Republic of Ireland (RoI) veterinary schools’ veterinary students typically undergo a five-year training programme, whereby they are trained in a variety of veterinary environments. These can be within university premises as well as on external placements. These are called extra-mural studies (EMS), and can be pre-clinical in nature (i.e. on farm, riding schools, kennels) or clinical (i.e. veterinary practices) (17, 18). It is unclear how these diverse environments might impact the risk of injury to veterinary students, or accident reporting.

Given these concerning data, and the paucity of contextual and behavioural analysis, further study is warranted. Considering that the main occupational hazard in the veterinary industry are animals, we decided to focus solely on animal-related injuries. Thus, the aim of this study was to establish the prevalence of animal-related injuries in UK and Irish veterinary students, to describe the context around these injuries, and to describe their consequences, both physically and behaviourally.

## Materials and methods

An online cross-sectional survey was designed. Respondents were asked about personal demographics (i.e. sex, age) and details of their university, year of study, and stage of course (ie pre-clinical or clinical). They were subsequently asked about animal-related injuries that they had acquired during their studies. This was followed by a series of more detailed questions about their most recent and their most severe injury. This included questions about where and how the injury was acquired, the species involved, medical consequences, student attitude to the injury, any resultant behaviour change, and the reporting process.

The survey was piloted with a small group of veterinary students and distributed through social media via accounts with large veterinary student followings, such as university veterinary societies. It was additionally distributed through email via official university webmail (Dublin, Edinburgh, Glasgow, Liverpool, and Nottingham). Any current veterinary student in the UK and Ireland was eligible to participate. The survey was open from July 8^th^ 2021 to August 31^st^ 2021.

Demographic characteristics of respondents were described. Overall injury prevalence was calculated and stratified by demographic data. Logistic regression was performed to identify any association between demographic variables and reporting an injury. Variables taken forward for multivariable analysis were selected through substantive knowledge and statistical significance. The analysis of questions related to the context of injury, attitude of injured person, and reporting culture were carried out descriptively, and stratified by the most prevalent species involved.

In the RoI, under The Safety, Health and Welfare at Work (Reporting of Accidents and Dangerous Occurrences) Regulations 2016 (S.I. No. 370 of 2016), employers legally must report to the Health and Safety Authority (HSA) any work-related injuries that result in an employee being unable to carry out normal work for more than three consecutive days (19). In the UK, under the Reporting of Injuries, Diseases and Dangerous Occurrences Regulations (RIDDOR), employees must report any ‘specified injury’ (which includes; fractures, amputations, loss of consciousness, amongst others), or if they are unable to work for more than seven days to the Health and Safety Executive (HSE) (20). However, as veterinary students are not employees there is no legal requirement to report injuries to the HSE or HSA nor to record time off studies. Thus, we have classified respondent reported injuries as RIDDOR reportable based solely on the list of specified injuries. The overall prevalence of these types of injuries was calculated and stratified by species.

Free text responses were analysed using an inductive content analysis (21, 22). Each response was initially read through, whilst notes around initial impression of content were created. Then, responses were considered in isolation and, labelled with a code that reflected these categories. Codes were created in an iterative fashion, and so were combined, revised, and deleted as more responses were read. The final set of codes were generated by repetition of this process to ensure internal validity of content codes. These were then counted to facilitate a quantitative comparison of content. Due to the nature of this intended analysis, responses to each question were coded separately. However, overall themes were then compared across different questions and across species.

Data masking was used to protect personally identifiable information of students; this was performed through aggregation of data or redacting of quotes. The study received ethical approval from the University of Liverpool Veterinary Research Ethics Committee (VREC1103).

## Results

There were 578 respondents to the survey; however, 7.8% (n=45) responses were excluded from analysis as 29 individuals had incomplete demographic information, and 16 individuals answered no questions past the demographics section. This resulted in 533 survey responses available for analysis. The estimated veterinary student population in 2021 in the UK and RoI was 7241. If all students were exposed to the survey, then the crude response rate was 7.4%. Respondents were predominately female, 18-24 years old, of white ethnicity, British, and with no self-identified disability (Table 1). These same demographics were reflected in those that had been injured. Almost 80% of respondents were from five of the ten universities where veterinary degrees could be obtained at the time of study. First year students were under-represented.

**Table 1.**
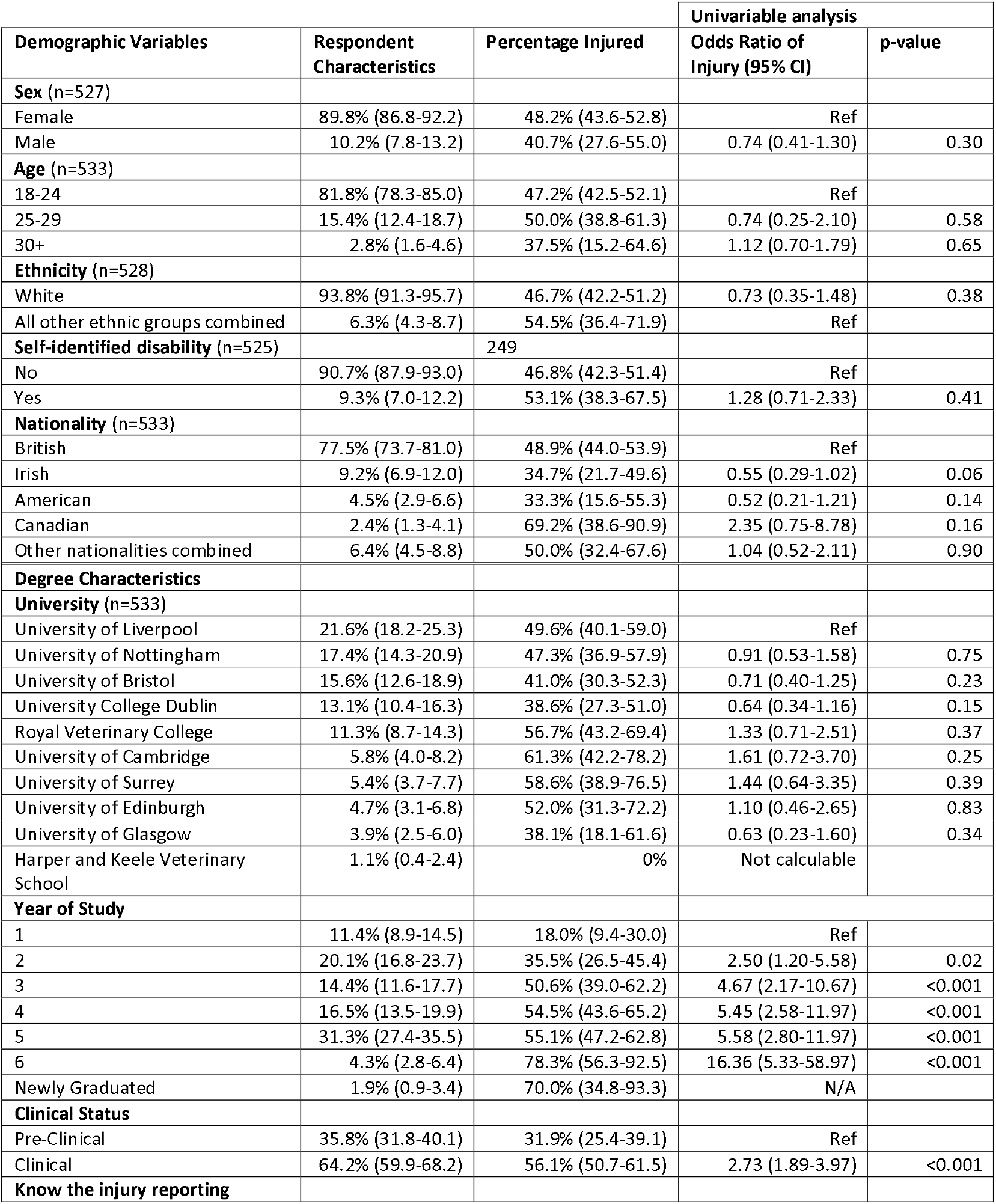
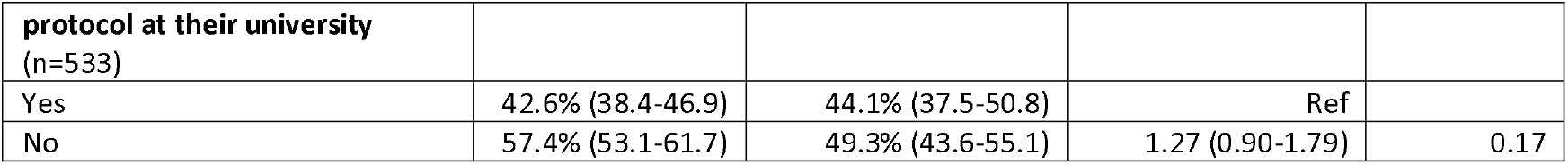
Demographics of veterinary students receiving animal-related injuries, and univariable analysis exploring demographic factors associated with acquiring such an injury.

Gender was excluded from analysis as there were less than 10 individuals that identified as being non-cis-gendered and we did not want these individuals to be personally identifiable. All ethnic groups, except the white British group, were aggregated into the group ‘all other ethnic groups combined’ as there were less than 10 individuals in these groups. Twenty-nine nationalities were represented, but 25 of these had less than 10 individuals and so these were aggregated into the group ‘other nationalities combined’. When asked about their academic year ten individuals wrote in the free text ‘New Graduate’, as the survey was distributed just after many students’ graduations, we included them for descriptive analysis as their experiences would relate to their time in veterinary education. However, they were excluded from the logistic regression.

Univariable analysis revealed that there was no association between any demographic variable or attendance of any one university and the odds of acquiring an injury (Table 1). The only significant associations were between year, and ‘stage of the degree’, and injury acquisition. The longer a student had been on the course the greater the odds of being injured, and students in their clinical years were more likely to have been injured than if they were in the preclinical part of the course. Due to strong collinearity (r=0.85) between the two univariable variables of interest (‘Year of Degree’ and ‘Stage of Degree’), a multivariable model was not created.

Overall, 47.5% (95% CI 43.2-51.8, n=253) of students reported having been injured by an animal during the veterinary degree, 35.5% (95%CI 31.4-39.7) of students reported being injured within the last 12 months. Of those injured, 71.1% (95% CI 65.1-76.7) reported having 1-3 injuries, 18.2% (95% CI 13.6-23.5) 4-5 injuries, 5.9% (95% CI 3.4-9.6) 6-9 injuries, and 3.6% (95%CI 1.6-6.6) having 10 or more injuries.

Ninety-five percent of responses (n=233/245) had enough contextual information to conclude whether their most recent injury was reportable to HSE via RIDDOR. Five percent of recent injuries (5.2%, 95%CI 2.7-8.8) were RIDDOR reportable. All these injuries were fractures, bar one concussion. Ninety-nine percent of responses (n=195/196) had enough contextual information to conclude whether their most severe injury was RIDDOR reportable. Nine percent of severe injuries (9.2%, 95%CI 5.6-14.2) were RIDDOR reportable. All these injuries were fractures or concussions.

Seventeen different species were involved in the students’ most recent injuries, and sixteen in their most severe injuries. Most recent injuries were caused by: companion animals (38.0%, 95%CI 31.8-44.6), livestock (37.6%, 95%CI 31.4-44.2), equids (23.5%, 95%CI 18.2-29.5), and wild animals (0.9%, 95%CI 0.1-3.1), with 87.8% linked to five species; horses, cattle, cats, dogs and sheep. For the most severe injuries, 48.7% (95%CI 41.5-56.0) involved livestock, 28.7% (95%CI 22.5-35.6) companion animals, 22.1% (95%CI 16.4-28.5) equids, and 0.5% (95%CI 0.0-2.8) wild animals, with 90.3% associated with the same five species.

Fifty percent (50.2%) of the most recent injuries occurred during the clinical part of the veterinary degree, 82.0% of injuries occurred on EMS placements, 15% of injured students sought medical treatment beyond first aid, 18.8% took more than two weeks to physically recover, but the majority (99.0%) did not take any time off their studies to recover. Only 16.8% of students informed their veterinary school about the injury. In contrast, 38.7% of the most severe injuries occurred during the clinical part of the veterinary degree, 87.8% of injuries occurred on EMS placements, 18.1% of injured students sought medical treatment beyond first aid, 26.9% took more than two weeks to physically recover, and 98.4% took no time off their studies to recover. Only 12.8% of students informed their veterinary school about the injury.

### Livestock-related Injuries

Over a third of the most recent injuries, and over 40% of the most severe injuries were associated with cattle and sheep. These overwhelmingly occurred in pre-clinical years, and on farm EMS placements (Table 2). Cattle-related injuries predominately occurred whilst the students were involved in three key activities; drug administration (23.3% of recent cattle injuries, and 24.2% of most severe), herding (21.4% and 15.2%), and milking (21.4% and 21.2%). More than one in five of these injuries occurred when the student was by themselves. Injuries were mainly the result of kicks (48.9% of recent, 47.1% of severe), and crushes (36.2% and 43.1%). The distribution of body parts injured was dispersed: hand (23.9% and 25.5%), leg (26.1% and 27.4%), arm (17.4% and 15.7%), foot (14.6% and 15.7%), and head (4.3% and 5.9%). The injuries were overwhelmingly bruises (76.6% and 68.6%), but fractures (8.5% and 11.8%) and concussion (2.0% of severe injuries) occurred. Six percent (6.4%) of recent injuries and 9.8% of severe injuries were RIDDOR reportable injuries.

**Table 2.**
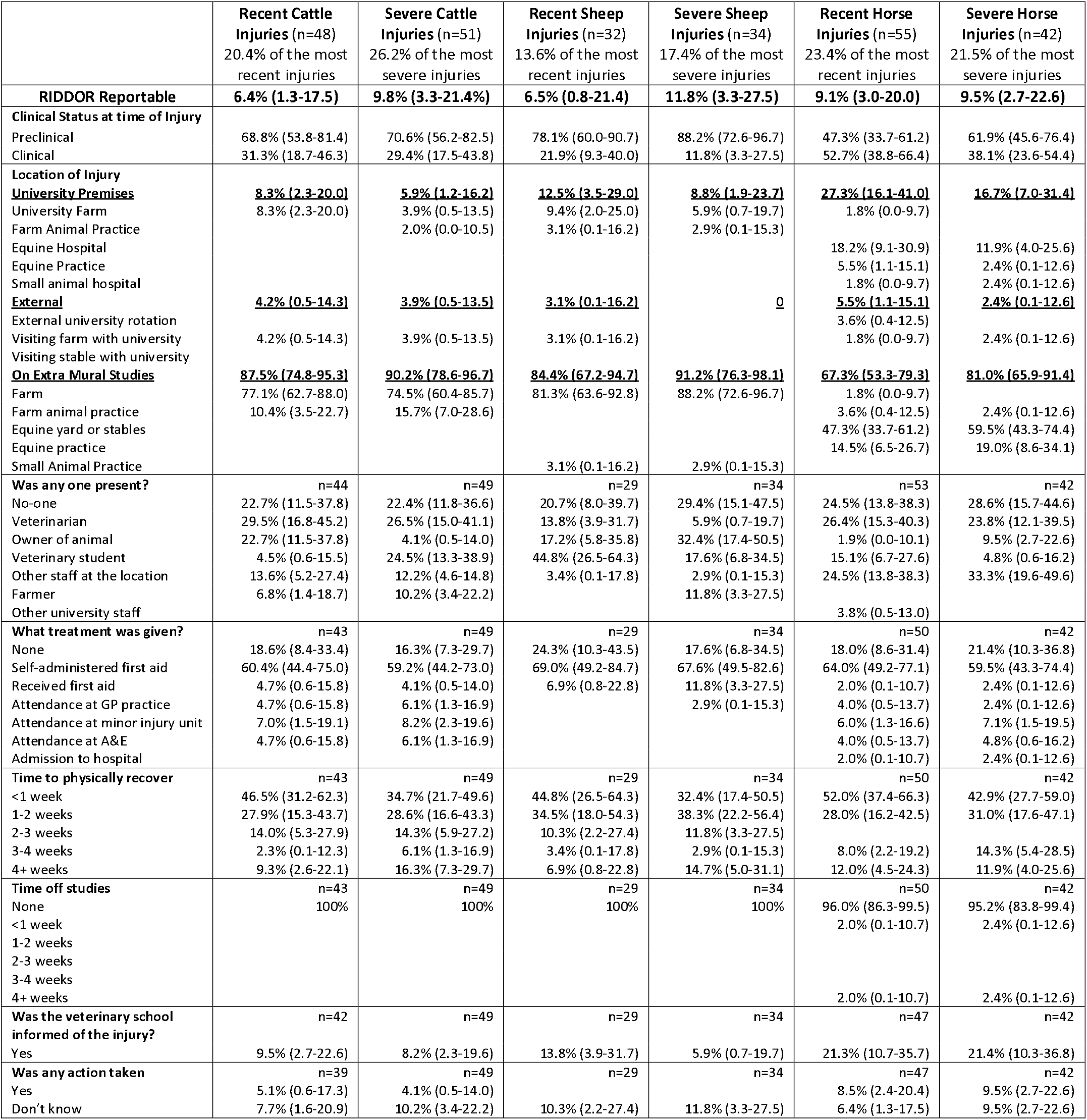
Context of injuries to veterinary students caused by livestock and horses.

Less than 20% of students received no medical treatment for their injuries, more than 60% had first aid treatment. Almost 12% (11.7%) of recent, and 14.3% of severe injuries required hospital treatment. Despite 25.6% of recent, and 36.7% of severe injuries taking more than 2 weeks to physically recover, no students took time off their studies. Less than 10% of students informed their university about their injury. Only two actions subsequent of reporting were recorded, these were one farm and one university investigation.

Sheep-related injuries predominately occurred whilst the students were involved in three key activities; restraint (55.2% of recent sheep injuries, and 41.7% of most severe), lambing (13.8% and 16.7%), and drug administration (13.8% and 12.5%). More than one in five of these injuries occurred when the student was by themselves. Injuries were mainly the result of crushes (25.8% of recent, 23.5% of severe), headbutts (22.6% and 20.6%), and being knocked over (9.7% and 14.7%). Injuries were focused on hands (51.6% and 41.2%) and legs (19.4% and 17.6), but a concerning number of head injuries occurred (9.7% and 17.6%). The main injuries were bruising (51.6% and 44.1%) and lacerations (12.9% and 17.6%), however there were concussions (3.2% and 5.9%), and 5.9% of most severe injuries were fractures. Seven percent (6.5%) of recent injuries and 11.8% of severe injuries were RIDDOR reportable injuries.

All head injuries were a result of being knocked over or head butted. These two quotes typify how these injuries were sustained:

> ‘*I was trying to catch a ewe and when I finally caught hold of it, it pulled me head first into a steel girder*.’
>
> ‘*I was trying to catch a ewe for lambing and as I grabbed her she threw her head up and hit me in the face*’

Over 70% of students had first aid treatment, and no students attended a hospital or minor injury unit. Despite 20.6% of recent, and 29.4% of severe injuries taking more than 2 weeks to physically recover, no students took time off their studies. Less than 15% of students informed their university about their injury. No students reported that any action was taken by a university resultant of their injury.

### Horse-related injuries

Around a quarter of the most recent injuries, and over 20% of the most severe injuries were associated with horses. The most recent injuries were evenly split between clinical and preclinical years, with the most common locations of injury being on an equine yard (47.3%), the university equine hospital (18.2%) and an EMS equine practice (14.5%) (Table 2). In contrast, the majority of the severe injuries occurred in preclinical years, and primarily occurred on equine yards (59.5%) and EMS equine practice (19.0%). Injuries occurred whilst the students were involved in three key activities; restraining a horse (30.2% of recent injuries and 22.9% of severe injuries), during a clinical examination or procedure (17.0% and 20.0%) and whilst leading a horse (13.2% and 20.0%).

The majority of the restraining-based injuries were for when a veterinarian was performing a procedure on the horse, for example:

> ‘*I was holding a young horse with a headcollar and lead rope while a vet administered an IM injection. The horse spooked at the injection and barged into me pressing me against the stable wall as it went through the stable door*.’

All clinical examination and procedure-based injuries were when the student was examining, or holding the X-ray plate by, a distal limb.

> ‘*After showing where the horse where our hands were, I inserted the X-ray plate medial to the stifle and was kicked on the leg*’

Around a quarter of all equine-related injuries occurred when the student was by themselves. Injuries were mainly the result of kicks (32.7% of recent, 40.5% of severe), bites (25.5% and 11.9%), and crushes (23.6% and 23.8%). The distribution of body parts injured was dispersed: leg (36.4% and 38.1%), arm (16.4% and 9.5%), foot (14.5% and 11.9%), hand (10.9% and 16.7%). Heads were involved in 3.6% and 7.1% of injuries, most of these were the result of kicks. Injuries were overwhelming bruises (74.5% and 69.0%), but there were fractures (10.9% and 11.9%) and dislocations (5.6% and 2.4%). Nine percent (9.1%) of recent injuries and 9.5% of severe injuries were RIDDOR reportable injuries.

Around one in five students received no treatment with the majority self-administering first aid. Twelve percent of recent and 14.3% of severe injuries required hospital treatment. Despite 20.0% of recent, and 26.2% of severe injuries taking more than 2 weeks to physically recover, only a very small percentage of students took time off their studies. Around one in five students informed their university about their injury. Only four actions or behaviour changes subsequent to reporting an incident were recorded. These included warning signs provided for specific horses, new pathways developed from yards to paddocks, and a referral to disability services.

### Companion animal

Over a third of the most recent injuries, and a quarter of the most severe injuries were associated with cats and dogs. These overwhelmingly occurred in clinical years, except for severe injuries caused by dogs where there was an even split between pre-clinical and clinical years (Table 3). The majority of injuries occurred on EMS placements, in particular in small animal practices. Cat-related injuries predominately occurred whilst students were involved in four key activities; restraint (51.3% of recent injuries and 50.0% of severe injuries), clinical examinations (10.3% and 8.3%), taking a cat in or out of a kennel (10.3% of recent injuries), and administering oral medication (7.7% and 12.5%). The majority of the time a veterinarian or veterinary nurse was present when the student was injured. Injuries were mainly the result of scratches (55.3% and 62.1%) and bites (44.7% and 37.9%). Three key body parts were injured; hands (46.8% and 51.7%), arms (44.7% and 37.9%) and heads (6.4% and 10.3%). The injuries were lacerations (53.2% and 51.7%) and puncture wounds (38.3% and 37.9%). There were no fractures or concussions. No cat-related injuries were RIDDOR reportable injuries.

**Table 3.**
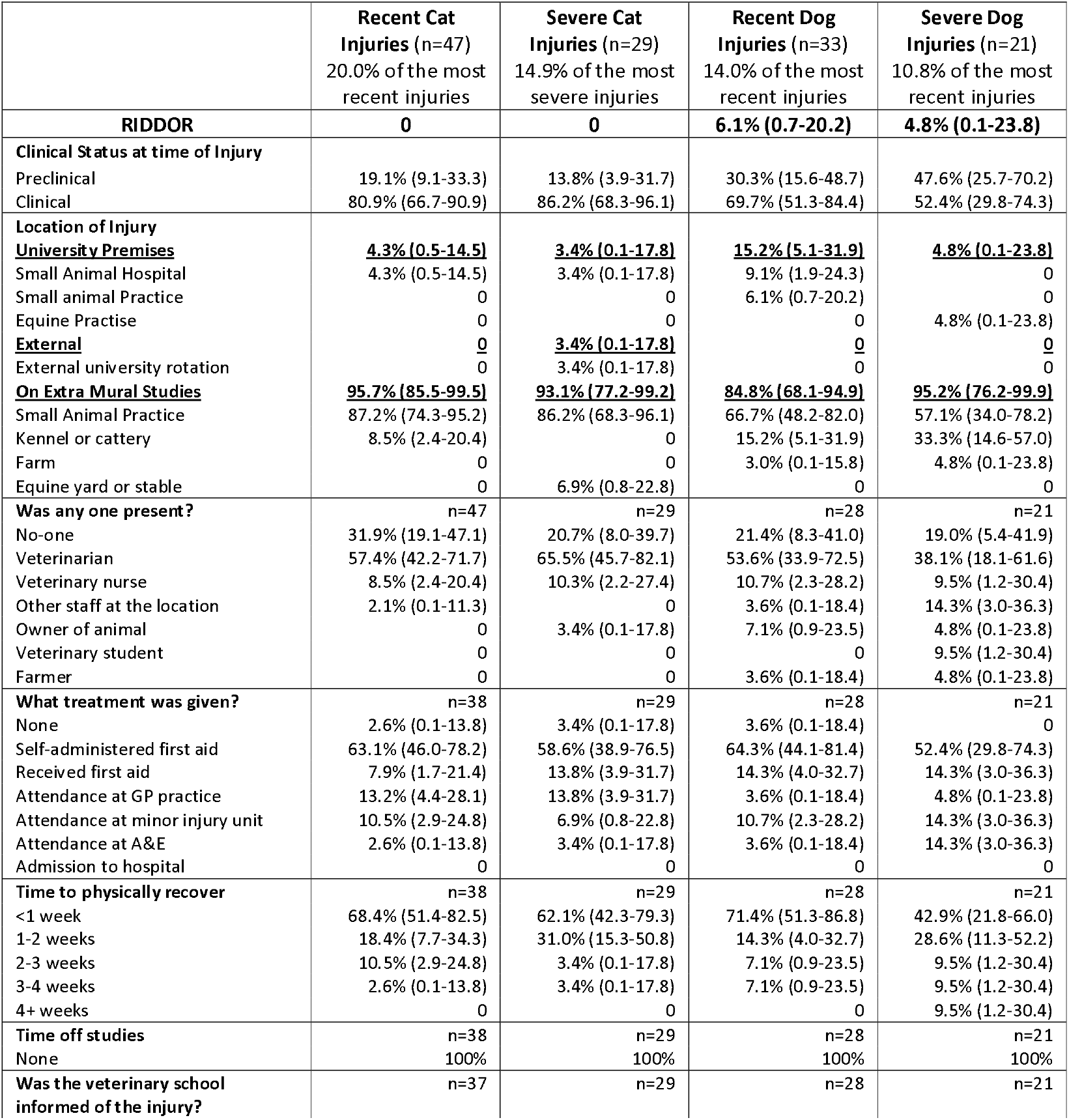
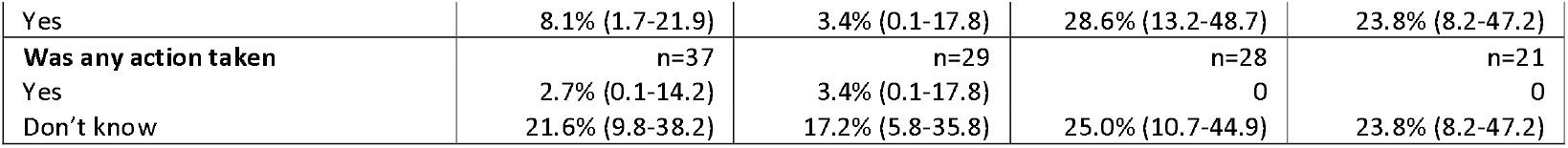
Context of injuries to veterinary students caused by companion animals.

The majority of students received medical treatment for their injuries, with more than 60% receiving first aid treatment. Thirteen percent (13.1%) of recent, and 10.3% of severe injuries required hospital treatment. Despite 13.2% of recent, and 6.8% of severe injuries taking more than 2 weeks to physically recover, no students took time off their studies. Fewer than 10% of students reported cat-related injuries to their university, compared with about one quarter for dog-related injuries.

Students were injured by dogs during many diverse activities, however 41.3% of recent injuries and 43.8% of severe injuries occurred when restraining a dog. The majority of the time a veterinarian or veterinary nurse was present when the student was injured. Injuries were mainly the result of bites (54.5% and 71.4%), scratches (27.3% and 9.5%) and head butts (9.5% of severe injuries). Three key body parts were injured; hands (57.6% and 66.7%), arms (27.3% and 19.0%), and legs (9.1and 9.5%). The injuries were lacerations (27.3% and 28.6%) and puncture wounds (30.3% and 28.6%) and bruising (21.2% and 23.8%). There were three fractures and no concussions. Six percent (6.1%) of recent dog-related injuries, and 4.8% of severe injuries, were RIDDOR reportable injuries.

Less than 4% of students received no medical treatment for their injuries, more than half had first aid treatment. Fourteen percent (14.3%) of recent, and 28.3% of severe injuries required hospital treatment. Despite 14.2% of recent, and 28.5% of severe injuries taking more than 2 weeks to physically recover, no students took time off their studies. Around of quarter of students informed their university about their injury. No actions were taken subsequent of reporting.

### Needlestick injuries

Almost five percent of recent injuries (4.7%, 95%CI 2.4-8.3) and 3.6% (95%CI 1.5-7.3) of the severe injuries were needlestick injuries. The main mechanisms involved were restraining the animal for a blood sample or IV catheter placement (27.8%), intra-muscular injection (22.2%), recapping a needle (16.7%), and having a needle in the pocket (11.1%). In only one instance was the drug in the syringe mentioned, namely pentobarbital.

### Qualitative content analysis

The majority of students did not experience emotional or mental effects resultant of their injuries (Table 4). When effects were mentioned the most prevalent themes were an increased anxiety or fear around a specific species and being more cautious. For example:

**Table 4.** Content analysis of behaviours reported by veterinary students subsequent to an animal-related injury.

| Themes | Cattle |  | Sheep |  | Horse |  | Cat |  | Dog |  |
| --- | --- | --- | --- | --- | --- | --- | --- | --- | --- | --- |
|  | Recent | Severe | Recent | Severe | Recent | Severe | Recent | Severe | Recent | Severe |
| Did you experience any **emotional or mental** effects resultant of the injury? | N=43 | N=49 | N=29 | N=34 | N=50 | N=42 | N=38 | N=29 | N=28 | N=21 |
| None | 62.8% | 63.3% | 55.2% | 73.5% | 62.0% | 59.5% | 78.9% | 82.3% | 78.6% | 71.4% |
| Increased anxiety or fear | 20.9% | 20.4% | 6.9% | 5.9% | 18.0% | 19.0% | 7.9% | 6.9% | 14.3% | 28.6% |
| More cautious around the animal | 2.3% | 6.1% | 13.8% | 2.9% | 12.0% | 14.3% | 7.9% | 6.9% | 3.6% | 0 |
| Won't work with that species again | 7.0% | 4.1% | 0 | 2.9% | 2.0% | 2.4% | 2.6% | 0 | 0 | 0 |
| Confidence knocked | 4.7% | 4.1% | 0 | 0 | 2.0% | 4.8% | 0 | 0 | 3.6% | 4.8% |
| Embarrassment | 2.3% | 2.0% | 6.9% | 2.9% | 4.0% | 2.4% | 2.6% | 3.4% | 0 | 0 |
| 'It made me stronger' | 4.7% | 2.0% | 0 | 5.9% | 2.0% | 2.4% | 0 | 0 | 0 | 0 |
| Why did you **not take time off** from your studies? | N=42 | N=49 | N=29 | N=34 | N=44 | N=40 | N=37 | N=28 | N=28 | N=21 |
| A minor injury | 61.9% | 57.1% | 75.9% | 76.5% | 59.1% | 52.5% | 78.4% | 82.1% | 75.0% | 61.2% |
| Felt no need as during their EMS period | 11.9% | 16.3% | 13.8% | 8.8% | 13.6% | 17.5% | 10.8% | 3.6% | 17.9% | 33.3% |
| Tight timetabling | 14.3% | 20.4% | 3.4% | 11.8% | 18.2% | 17.5% | 2.7% | 0 | 0 | 0 |
| Could still function, just! | 11.9% | 8.5% | 3.4% | 0 | 11.4% | 15.0% | 5.4% | 10.7% | 3.6% | 4.8% |
| Didn't want to get in trouble with university or EMS provider | 7.1% | 4.1% | 6.9% | 2.9% | 15.9% | 15.0% | 0 | 3.6% | 0 | 0 |
| 'Just get on with it!' | 7.1% | 4.1% | 0 | 2.9% | 4.5% | 10.0% | 2.7% | 3.6% | 3.6% | 0 |
| Why did you choose **not to report** the incident? | N=36 | N=42 | N=25 | N=32 | N=38 | N=40 | N=24 | N=19 | N=19 | N=21 |
| Only minor/wasn't serious enough | 61.1% | 47.6% | 68.0% | 59.4% | 65.8% | 52.5% | 79.2% | 78.9% | 68.4% | 33.3% |
| Unaware needed reporting | 13.9% | 23.8% | 16.0% | 25.0% | 15.8% | 17.5% | 20.8% | 21.1% | 36.8% | 28.6% |
| Didn't want to make a fuss | 13.9% | 19.0% | 0 | 0 | 15.8% | 17.5% | 8.3% | 15.8% | 10.5% | 9.5% |
| Too much effort (what's the benefit?) | 16.7% | 26.2% | 8.0% | 12.5% | 10.5% | 15.0% | 4.2% | 0 | 5.3% | 4.8% |
| Felt silly | 8.3% | 7.1% | 12.0% | 6.3% | 5.3% | 15.0% | 8.3% | 10.5% | 0 | 4.8% |
| It's just one of those things/part of the course | 2.8% | 4.8% | 4.0% | 12.5% | 5.3% | 10.0% | 4.2% | 5.3% | 5.3% | 4.8% |
| Have you **changed your behaviour** around the animal species involved in the injury? | N=43 | N=39 | N=43 | N=34 | N=50 | N=42 | N=38 | N=29 | N=28 | N=21 |
| No | 32.6% | 41.0% | 37.2% | 55.9% | 44.0% | 47.6% | 44.7% | 37.9% | 39.3% | 28.6% |
| Caution/Less confident | 34.9% | 41.0% | 4.7% | 8.8% | 16.0% | 11.9% | 7.9% | 10.3% | 21.4% | 33.3% |
| Won't do procedure/avoid animal | 4.7% | 5.1% | 0 | 2.9% | 8.0% | 7.1% | 5.3% | 6.9% | 7.1% | 4.8% |
| Do procedure differently | 4.7% | 20.5% | 9.3% | 11.8% | 8.0% | 7.1% | 7.9% | 6.9% | 0 | 4.8% |
| Improve handling/restraint | 11.6% | 10.3% | 4.7% | 2.9% | 0 | 4.8% | 26.3% | 20.7% | 28.6% | 28.6% |
| Use personal protective equipment | 0 | 2.7% | 0 | 0 | 16.0% | 16.7% | 0 | 0 | 0 | 0 |
| Denial that did anything wrong | 9.3% | 5.1% | 7.0% | 2.9% | 2.0% | 0 | 5.3% | 6.9% | 7.1% | 9.5% |
| Improved awareness of surroundings | 9.3% | 17.9% | 4.7% | 14.7% | 8.0% | 9.5% | 0 | 0 | 0 | 0 |

> ‘*Fear of getting kicked by cows and anxiety around cows in general’-Fractured hand, cow kick ‘Fear of being bitten again*’ – Hand laceration, dog bite

Some livestock- and horse-related injuries led students to want to avoid these species professionally;

> “*I only know I won’t be working with cows, thank you very much*” – Bruised hand, cow crush

> “*Less confidence with horses. I already wasn’t equine inclined but I am experienced with handling and riding horses. However this experience made me realise how dangerous horses can be especially in a veterinary context and it has made me more nervous to give horses injections. It has definitely confirmed I don’t want to work in equine practice*.” – Bruised arm, horse crush

Others stated that their confidence was knocked, and that becoming injured was a source of embarrassment;

> “*Embarrassment for getting upset in front of the vet (got teary due to the shock of the situation at the time not due to pain*)” – Face laceration, cat scratch

> “*I got very bad feedback from the EMS provider, I believe they were probably scared of the fact I was hurt so wanted to make it seem as though I was incompetent. This affected my confidence for at least a year*” – Hand laceration, dog bite

Surprisingly some students, though only those injured by livestock or horses, stated that the injuries made them stronger and were beneficial to them:

> “*It made me stronger. Bumps and bruises are part of the working farm environment, even if you get bowled over once in a while*.” – Bruised leg, cow trampling

The main reason that the majority of students gave for not taking time off their studies was that they had only received a minor injury. Based on the injury description this was accurate for most cases. A proportion did not view EMS as part of their studies, and so time off from EMS was not viewed as time off their studies to recover.

> “*I attended placement in a holiday and so I was fully healed by the time I started back at uni*” – Laceration of arms and head, pig goring

Many, especially those injured by cattle and horses, felt they could not take time off as there was no resource allocation to reschedule their placements;

> “*I knew that if I took time off of the ems placement I’d have to resit it and I didn’t have the time or money as it cost me £40 a day to travel there*” – Bruised arm, cow kick

> “*I felt I had to sort of grin and bear it because I worry that you cant really take time off veterinary. We have so much EMS to fit in and attendance of our lectures/practicals is very closely monitored*.” – Abdominal bruising, multiple horse kicks

Some students, despite having relatively serious injuries, felt that they could still function enough to work and so did not take time off;

> “*Didn’t inhibit me too much and I’ve broken fingers before so didn’t bother me too much*” – Fractured hand and fingers, cattle crush

Some, especially those injured by horses and livestock, felt that the injury could get them in trouble with either their university or EMS provider;

> “*Was told by host practice if I left the placement to recover I would not be welcome to return*” – Fractured skull, horse kick

> “*I needed to complete the EMS placement. I was worried my supervisor wouldn’t have signed my placement if I took a time off*” – Bruised abdomen, trampled by horse

Many students described feeling that they needed to be tough and ‘get on with it’;

> “*You just have to get on with it. A bruise or a cut shouldn’t stop you from being able to do what you have to*” – Bruised head, sheep head butt

> “*I was determined to push through and just deal with it, and I did*.” – Friction burn of hand, escaped horse

> “*Felt like I had to show I was tough and walk it off”- Sprained leg, knocked over by horse*

Most students who didn’t report injuries felt they weren’t serious enough, though perceptions of “severity” were influenced by reporting concerns and an expectation of injury:

> “*it was not serious, I would not want a report to impact my studies or the practice I was at just because a cat had good aim. I recognize that as an everyday hazard of the job*.” – Bruised hand, cat scratch

Many described being unaware that reporting was needed. Some did not want to “make a fuss”, whilst others felt it was too onerous and could not see any benefit to reporting, compared with the perceived negative impact of “making a fuss”.

> “*Can’t be arsed with paperwork, seems a bit dramatic, I didn’t lose a finger. I’d probably be laughed at and they wouldn’t know how to report. The outcome wouldn’t benefit me so why would I*?” – Puncture wound to hand, fox bite

> “*Not necessary - protocol is idiotic*” – Arm pain, dog bite

> “*What difference would it make other than making me fill out paperwork*” – Jaw fracture, horse kick

Some felt there would be personal emotional repercussions of being made to feel stupid, embarrassed, or not taken seriously.

> “*I didn’t want them to force me to take time off*” – Foot fracture, cow crush
>
> “*It was a one-vet, one-nurse practice. This vet is very unprofessional and said to “grow up, it happens” and barely gave me the time to remove the nail (which was hanging off) and wash & bandage it before asking for help with the next animal. The nurse was too afraid to speak up in front of the vet*.” – Fingernail avulsion, dog headbutt

Worryingly, many felt that injuries were just part and parcel of being a veterinarian and so they didn’t need reporting:

> “*It was only a bruise and a bit of an occupational hazard*!” – Leg bruise, sheep kick
>
> “*Risks of doing TPR, everyone gets kicked at some point*” – Leg bruise, horse kick

More than a third of students stated that they would not change their behaviour around the species involved in their injury, and others stated that the injury was inevitable and could not have been avoided.

> “*No, I was just unlucky*” – Leg bruise, cow kick

Of those that did say they would change their behaviour, many said that they would exercise caution or be less confident around those animals, whilst other would use greater restraint, be more aware of their surroundings, do the procedure differently, and wear PPE.

> “*More cautious around handling dogs, will muzzle dogs more often*” – Leg laceration, dog bite
>
> “*Would perhaps be more likely to wear a helmet around the yard as well as riding. It could easily have been my head trampled instead of my legs*”. - Sprained leg, knocked over by horse.

> “*Will refuse to work with cows if there is no proper restraint From now on, crush or nothing*” – Bruised hand, cow crush

Some individuals stated that they would just avoid that species in their future careers, suggesting that perceived risk of injury might be key in shaping the veterinary workforce.

> “*There was nothing I could have done to avoid the injury so there is nothing I can change other than just not working with them*” – Bruised arm, cow kick

## Discussion

This study explores veterinary student injuries and resultant behaviours during formal training, an area that has received little prior attention. As expected, we found that levels of injury are higher than previous audits of veterinary school injury records (5), as students reported being unwilling to use formal reporting systems. We report a concerningly high level of injury, with almost half of students being injured by an animal during their degree, and an annual prevalence of around 35%. Many students experienced multiple injuries throughout their studies. The most severe injuries are occurring within pre-clinical years. Around 5% of students most recent injuries reported in this survey were sufficiently severe to be considered RIDDOR reportable.

This study finds that the risk of injury is highest on EMS placements. This finding is concerning but unsurprising, as students are in diverse placement types and locations, with variable safety standards and supervision. Qualitative data yields some concerning scenarios; for example, students being placed in risky situations where they felt unable to mitigate for potential harm; being improperly trained; and feeling that supervisors in those placements would think them “weak” if they mentioned or reported injuries. Power imbalances appear present, as students were marked on their placements and ‘making a fuss’ about injuries could be deemed to lower their marks. EMS placements provide a challenge to the veterinary schools as students are not under their direct supervision at this point. These placements are an integral part of the veterinary curriculum and highly beneficial for student learning and as such it is vital that students are given additional support during them in order to recognise and avoid risk, and to empower students to report placements where health and safety standards are low.

We found that accident reporting is low overall, with few students reporting their injuries. If one were to extrapolate out our findings to the whole student population, at least 2554 injuries would occur annually. An audit of five UK veterinary schools found on average 58 injuries reported annually (5). This highlights that currently reporting systems do not capture the full extent of the injuries that students are experiencing, particularly on EMS placements. This underestimation will inhibit universities’ abilities to make meaningful changes to student safety and thus warrants attention. However, whether students should report injuries to a veterinary school is a legal grey area. Students in the UK are not legally required to report every injury to their university, but institutions typically require reporting for incidents that occur on campus or during university-organised activities so they can meet their own health and safety legal obligations (19). Injuries that happen outside university activities (such as EMS) generally carry no obligation for students to report them. Many veterinary schools see that students are under their care and have a moral obligation to their safety when on placements and so encourage students to report accidents, incidents or near misses. Reasons that students gave for not reporting reflect those frequently found in the veterinary profession; lack of knowledge of reporting structures; lack of confidence in meaningful change as a result of reporting; and disinclination to draw attention to the event, sometimes for fear of being blamed (3, 4). It is unclear whether the students are mirroring or embracing the culture of the veterinary profession, or whether these behaviours are established before attending university. This warrants further exploration.

This culture of under-reporting is important because it could establish long-term habits that persist into professional practice, reducing the visibility of workplace risks and limiting prevention opportunities. In other industries, effective interventions which encourage reporting have focussed on improved dialogue and feedback between accident reporters and management, with visible improvements to systems on the basis of reports (23-25). Applying a similar approach at veterinary schools could foster a culture that encourages transparent reporting, whilst maintaining anonymity, and one that emphasises the role that reporting has in improving student safety and animal handling practices.

Few students sought medical treatment beyond self-administered first aid or took time off their studies to recover; even when the injury was severe enough to warrant these behaviours. Students often attributed this to beliefs that injury was an expected part of veterinary training, that “toughness” was valued, and to limited resources (e.g., time or funding) to pause studies or repeat placements. This appears particularly relevant for younger individuals, who may feel their input has limited influence on safety improvements, which is especially common among young women in other workplaces (26). Females make up the majority of veterinary school students and thus exploring the impacts of gender on injury prevalence, perception, and the need to display “toughness” in future studies could yield valuable insight. The culture of limited injury care and limited time off work reflects very closely that of the veterinary profession (3, 4). This is shaped by heavy workloads and academic demands, which could discourage rest or recovery, as well as by a culture of stoicism and presenteeism that normalises working through pain or injury. Over time, this normalisation of risk could reinforce underreporting and underappreciating the seriousness of injuries, embedding habits that students are likely to carry into their professional careers.

The prevalent events preceding cattle-related injuries (drug administration, herding, and milking) were primarily reflective of the students’ activities during pre-clinical EMS. From the responses it is unclear why injuries during these events were more prevalent and further exploration is needed. The injuries received were relatively less traumatic than what is reported in the literature, which predominately focuses on individuals attending emergency departments or admissions to hospital for cattle related injuries (less than 10% of students within our data) (27-31). This is likely due to the lack of studies describing cattle-related injuries not requiring hospital treatment. Sheep-related injuries were reflective of common activities performed during EMS placements (ie restraint, lambing, drug administration). Leg and head injuries typically resulted from being headbutted or knocked over, leading to severe outcomes such as concussions and fractures. Similarly, cattle-related injuries resulted in fractures (6%) and concussions (10%). For both cattle- and sheep-related injuries over 20% of students were injured while alone. These findings highlight concerns around student safety and vulnerability during EMS placements, particularly given the risks of delayed treatment, and underscore the need to consider how best to safeguard students on farms.

Horse-related injuries occurred whilst the students were restraining or leading a horse, or when examining a distal limb. Kicks to legs and crushed feet were the main mechanisms of injury. However, they received more bites than anticipated, and fewer head injuries (2, 4). The bites are likely reflective of the frequency of close-hand tasks, such as restraint and leading horses, which compared to students, vets are less likely to perform. Conversely, vets are more likely to perform clinical tasks that increase the risk of a head injury, such as distal limb examinations and procedures, with students often positioned as handlers. When head injuries did occur, they were often during a distal limb examination. There were a number of fractures and dislocations, highlighting that equine EMS placements still pose a substantial risk of severe injury. Some students described subsequent efforts to better understand horse behaviour, recognising that improved awareness of equine body language may help anticipate and prevent risky situations. While reflection is valuable for personal learning, reliance on individual adaptation alone risks placing responsibility on students rather than addressing systemic safety issues. Embedding reflection within a wider safety culture, supported by structured training, supervision, and institutional commitment, would ensure that both individual and organisational factors contribute to safer practices during veterinary education and future clinical work.

Cat-related injuries were most commonly sustained during restraint, clinical examination, taking a cat in/out of a kennel, and administration of oral medications. These are all routine tasks that a student would undertake at a university or on an EMS placement. These incidents typically occurred in the presence of a veterinarian or nurse, suggesting that even supervised settings can carry risks. While injuries were generally less severe than those associated with large animals, with no fractures or concussions reported, cat bites frequently resulted in puncture wounds to the hands and wrists. Such injuries are clinically significant, as cat bites can rapidly lead to serious infections, and current recommendations state that all cat bites should be assessed at a hospital and treated with antibiotics (32). However, only around one in ten students sought hospital care following a bite. This raises concerns about whether students were unaware of the potential risks of a cat bite or were consciously choosing to ignore them, reflecting gaps in both education and safety culture around the management of small animal-related injuries. Students’ reluctance to seek care may reflect a professional culture of ‘just getting on with it’ and minimising so-called minor injuries, norms that are systemic within the veterinary profession (3, 14). Such attitudes may indicate that the prevailing industry culture is shaping students’ behaviours, normalising risk and potentially discouraging the adoption of practices that prioritise personal health and safety. This underscores the need to address cultural factors within veterinary training to foster a safety-oriented mindset from the outset of professional practice

Injuries caused by dogs showed considerable contextual variation, with no clear trends across activities. The majority of incidents involved bites, reflecting the well-documented risk dogs pose in veterinary practice. Unlike previous research that has highlighted a notable proportion of dog bites to heads (3, 5), this study found relatively few such cases. This discrepancy may be explained by the supervised nature of student involvement, where students are often tasked with handling or restraint rather than performing procedures that place the head and upper body near the animal. The variation observed highlights the unpredictable nature of canine behaviour and the multiple points of contact between students and dogs during clinical training. It also underlines the importance of embedding robust safety practices and animal-handling training early in the veterinary curriculum.

The qualitative content analysis revealed the extent to which culture plays a role in accident experience, behaviour, and reporting. It was clear that students expected injuries to occur as part of their training, including severe injuries. As such, they described feeling a need to limit reporting and avoid being seen to “make a fuss”, in the face of placement providers and other veterinarians. This may be at least partly caused by the tendency toward blaming the injured party, as well as the fear of having marks downgraded. The resulting culture of bravado around injury has the potential to increase levels of harm, for example by students feeling that they are implicitly expected to put themselves in risky situations, and by reducing help-seeking behaviour after incidents occur. Research examining injury risk factors and strategies for improving safety emphasises the need for cultural change, including shifts in managerial practices and organisational norms (33-35). Given that graduate veterinary surgeons have been shown to accept injury as an inherent part of their work, and that the likelihood of injury is influenced by local workplace culture, it is essential to examine how culture operates across the profession.

The identification of a poor safety culture in the veterinary student population should be of serious concern for the profession. We do not know where this culture begins, whether before or during veterinary education, on placement or at university sites. Some students may already carry attitudes shaped by prior experiences with animals, such as work-experience, where risk-taking or “getting the job done” was implicitly rewarded. Once at vet school, these attitudes could be reinforced if safety protocols are inconsistently enforced or if students witness peers or instructors normalising minor injuries or risky practices. EMS placements may further expose students to high-pressure and fast-paced environments, where time constraints, hierarchical dynamics, and the prioritisation of patient care over personal safety can encourage shortcuts or silence around reporting injuries. Collectively, these experiences may influence how students come to understand which behaviours are tolerated, which risks seem acceptable, and whose voices are heard in discussions about safety. If these patterns persist, they could contribute to longer-term habits and professional norms that students carry into their veterinary careers. Further research is needed to understand whether and how such early experiences might shape a culture in which safety is undervalued, and preventable harm becomes routine.

This study has limitations, particularly including the use of the survey methodology bringing issues of reporting bias. For example, students may have felt more compelled to respond to the survey if they had experienced an injury or accident during their studies, even though steps were taken to mitigate this risk (the advertisement was designed to encourage participation for those who both had and hadn’t experienced injury or illness). Reassuringly, the demographics of participants reflected that of veterinary students in the UK, suggesting a broadly representative sample.

## Conclusions

This survey provides valuable insights for the veterinary profession into the realities students face around injury and injury-related behaviours during their training. We identify concerningly high levels of injury and severe injury, much of which is under-reported, alongside a culture of injury acceptance and expectation. The activity-specific risks highlighted in our data should assist universities and placement providers to strengthen, training, support and reporting structures. However, the expectation of injury and the tendency to continue working while injured remain particular concerns, and organisations may benefit from drawing on lessons from other sectors that have successfully shifted cultures around risk management and reporting.

## Ethics Statement

The study received ethical approval from the University of Liverpool Veterinary Research Ethics Committee (VREC1103).

## Author’s contributions

JT, TF: Conceptualisation, Methodology, Analysis, Writing – Review & Editing. LK: Methodology, Analysis, Writing – Original Draft. GP: Conceptualisation, Writing – Review & Editing

## Acknowledgements

We would like to thank all the universities and social media groups that helped disseminate this survey, in particular ‘VetWings’.

## Data Availability

The raw data for this study are unlimited free text responses to survey questions, which contain potentially identifying and sensitive patient information. We are unable to share these data publicly because of restrictions by the University of Liverpool Veterinary Research Ethics Committee as participants did not consent to sharing of their data outside of the study team. Relevant, de-identified excerpts of the transcripts are included in the paper. Requests for additional information can be sent to the University of Liverpool Veterinary Research Ethics Committee at.

## References

1. Statistics UBoL,. Survey of Occupational Injuries and Illnesses Data 2023 [Available from: https://www.bls.gov/iif/nonfatal-injuries-and-illnesses-tables.htm.

2. Parkin TDH, Brown J, Macdonald EB. Occupational risks of working with horses: A questionnaire survey of equine veterinary surgeons. Equine Veterinary Education. 2018;30:200–5.

3. Tulloch JSP, Schofield I, Jackson R, Whiting M. ‘Just part of the job’ - understanding work-related injuries and safety culture in companion animal veterinary practices. J Small Anim Pract. 2025.

4. Tulloch JSP, Schofield I, Jackson R, Whiting M. ‘It’s only a flesh wound’ - Understanding the safety culture in equine, production animal and mixed veterinary practices. Prev Vet Med. 2025;241:106541.

5. Tulloch JSP, Fleming KM, Pinchbeck G, Forster J, Lowe W, Westgarth C. Audit of animal-related injuries at UK veterinary schools between 2009 and 2018. Vet Rec. 2023;193(7):e3171.

6. Johnson L, Fritschi L. Frequency of workplace incidents and injuries in veterinarians, veterinary nurses and veterinary students and measures to control these. Aust Vet J. 2024;102(9):431–9.

7. Fritschi L, Day L, Shirangi A, Robertson I, Lucas M, Vizard A. Injury in Australian veterinarians. Occup Med (Lond). 2006;56(3):199–203.

8. Gabel CL, Gerberich SG. Risk factors for injury among veterinarians. Epidemiology. 2002;13(1):80–6.

9. Epp T, Waldner C. Occupational health hazards in veterinary medicine: physical, psychological, and chemical hazards. Can Vet J. 2012;53(2):151–7.

10. Whittem T, Woodward AP, Hoppach M. A Survey of Injuries That Occurred in Veterinary Teaching Hospitals during 2017. J Vet Med Educ. 2021;48:401–16.

11. Trimpop R, Austin EJ, Kirkcaldy BD. Occupational and traffic accidents among veterinary surgeons. Stress Medicine. 2000;16:243–57.

12. Voss DS, Boyd MV, Evanson JF, Bender JB. An increase in animal-related occupational injuries at a veterinary medical center (2008-2022). J Am Vet Med Assoc. 2024;262(3):376–82.

13. Irwin A, Vikman J, Ellis H. ‘No-one knows where you are’: veterinary perceptions regarding safety and risk when alone and on-call. Vet Rec. 2019;185(23):728.

14. Furtado T, Whiting M, Schofield I, Jackson R, Tulloch JSP. Pain, inconvenience and blame: defining work-related injuries in the veterinary workplace. Occup Med (Lond). 2024;74(7):501–7.

15. Weaver DR, Newman LS, Lezotte DC, Morley PS. Perceptions regarding workplace hazards at a veterinary teaching hospital. J Am Vet Med Assoc. 2010;237(1):93–100.

16. Furtado T, Kennedy L, Pinchbeck G, Tulloch JSP. Zoonotic infections in UK and Irish veterinary students: a cross-sectional survey. BMC Public Health. 2024;24(1):1272.

17. RCVS,. Extra-mural studies (EMS) 2023 [Available from: https://www.rcvs.org.uk/lifelong-learning/students/veterinary-students/extra-mural-studies-ems/.

18. Medicine USoV,. Clinical Extramural Studies (CEMS) 2022 [Available from: https://www.ucd.ie/vetmed/study/clinicalextramuralstudies/.

19. Authority HaS,. Accident and Dangerous Occurence Reporting [Available from: https://www.hsa.ie/eng/topics/accident_and_dangerous_occurrence_reporting/accident_and_dangerous_occurrence_reporting.html#:~:text=Call%3A.

20. Executive HaS. Reporting of Injuries, Diseases and Dangerous Occurrences Regulations 2013 - RIDDOR - HSE 2013 [Available from: https://www.hse.gov.uk/pubns/indg453.pdf.

21. Krippendorff K,. Content Analysis: An Introduction to Its Methodology: SAGE Publications, Inc.; 2019.

22. Vears DF, Gillam L. Inductive content analysis: A guide for beginning qualitative researchers. Focus on Health Professionals. 2022;23:111–27.

23. Fox CJ, Sulzer-Azaroff B. Increasing completion of accident reports. Journal of Safety Research. 1987;18:65–71.

24. Hale AR, Guldenmund F, van Loenhout PLCH, Oh JIH,. Evaluating safety management and culture interventions to improve safety: Effective intervention strategies. Safety Science. 2010;48:1026–35.

25. Benn J, Koutantji M, Wallace L, Spurgeon P, Rejman M, Healey A, Vincent C. Feedback from incident reporting: information and action to improve patient safety. Qual Saf Health Care. 2009;18(1):11–21.

26. Curtis Breslin F, Polzer J, MacEachen E, Morrongiello B, Shannon H. Workplace injury or “part of the job”?: towards a gendered understanding of injuries and complaints among young workers. Soc Sci Med. 2007;64(4):782–93.

27. Rhind JH, Quinn D, Cosbey L, Mobley D, Britton I, Lim J. Cattle-Related Trauma: A 5-Year Retrospective Review in a Adult Major Trauma Center. J Emerg Trauma Shock. 2021;14(2):86–91.

28. Lucas M, Day L, Fritschi L. Serious injuries to Australian veterinarians working with cattle. Aust Vet J. 2013;91(1-2):57–60.

29. Ehrhard S, Botte F, Klukowska-Rotzler J, Exadaktylos AK, Jakob DA. Cattle-related trauma: a 10-year retrospective cohort study of patients admitted to a single tertiary trauma centre in Switzerland. Swiss Med Wkly. 2022;152:w30201.

30. Byard RW. Death and injuries caused by cattle: A forensic overview. Forensic Sci Med Pathol. 2025;21(1):401–5.

31. Savage G, Liesegang A, Campbell J, Lyon M, Fry D. Horse and Cattle-Related Trauma: A Retrospective Review of Injuries and Management in a Regional Queensland Hospital. Cureus. 2023;15(3):e35746.

32. Colmers-Gray IN, Tulloch JS, Dostaler G, Bai AD. Management of mammalian bites. BMJ. 2023;380:e071921.

33. Dyreborg J, Lipscomb HJ, Nielsen K, Torner M, Rasmussen K, Frydendall KB, et al. Safety interventions for the prevention of accidents at work: A systematic review. Campbell Syst Rev. 2022;18(2):e1234.

34. Probst TM, Brubaker TL, Barsotti A. Organizational injury rate underreporting: the moderating effect of organizational safety climate. J Appl Psychol. 2008;93(5):1147–54.

35. Probst TM, Estrada AX. Accident under-reporting among employees: testing the moderating influence of psychological safety climate and supervisor enforcement of safety practices. Accid Anal Prev. 2010;42(5):1438–44.

